# RELATIONSHIP BETWEEN THE LEVEL OF SPIRITUALITY AND BLOOD PRESSURE CONTROL AMONG ADULT HYPERTENSIVE PATIENTS IN RURAL SOUTHWESTERN NIGERIA

**DOI:** 10.1101/2023.03.08.23287012

**Authors:** Adetunji Omonijo, Paul Olowoyo, Azeez Oyemomi Ibrahim

## Abstract

**Background:** Spirituality has been strongly associated with good blood pressure control as it forms a strong coping mechanism in hypertensive patients.

**Purpose:** This hospital-based cross-sectional study was done to determine the relationship between spirituality and blood pressure control among adult hypertensive patients in rural Southwestern Nigeria with a view to achieving good blood pressure control.

**Method:** The selection was done by systematic random sampling technique. Socio-demographic and clinical information were obtained through semi-structured interviewer-administered questionnaires. The level of spirituality was assessed using the Spiritual Perspective Scale. Data were analysed using the Statistical Package for Social Sciences version 20.0. Statistical significance was set at p ≤ 0.05.

**Results:** The mean age of the respondents was 61.1±11.1 years. More than half (52.6%) had high level of spirituality and more than two-thirds (67.1%) of respondents had controlled blood pressure. Respondents with high level of spirituality were 4.76 times more likely to have a good blood pressure control {p<0.001, 95%CI (1.05-14.99)}than those with low level of spirituality.

**Conclusion:** Proper understanding and effective utilization of this relationship will assist health professionals and researchers in the appropriate integration of this concept into patients’ holistic care with the aim of achieving better blood pressure control among hypertensive patients.

## Introduction

Hypertension and its associated complications have become a common and important global public health challenge of the 21^st^ century.^1-3^ Hypertension is a major risk factor and a powerful predictor of cardiovascular morbidity and mortality.^1,4^

The global prevalence of hypertension has been increasing. For instance, in the year 2000, the world was estimated to have close to 1 billion people with hypertension with a prevalence rate of 26.4% and this has been projected to increase to 1.56 billion affected individuals with a prevalence rate of 29.2% in 2025, the population burden being greater in developing countries.^2,5^ In Africa, blood pressure control rates were uniformly low and often does not exceed 45% of the hypertensive population.^7^ An hospital-based study in Port Harcourt, Nigeria shows that good hypertension control could only be achieved for just 24.2% of the patients seen in the clinic.^6^ According to a community-based study among an urban population in Zaria, Nigeria shows that hypertension control rate was only 12.4% among patients who have been on antihypertensive medications for over 3months.^6^ These gloomy statistics call for urgent action, especially as the WHO has projected a further 24% increase in the prevalence of non-communicable diseases in Nigeria, in the next 10 years.^7^

Worldwide, there has been a systematic introduction of the concept of spirituality into the medical field with a growing interest in the possible health benefits as a result of having a spiritual belief and following a religious lifestyle.^8^

Spirituality can be defined as “the aspect of humanity that refers to the way individuals seek and express meaning and purpose and the way they experience their connectedness to the moment, to self, to others, to nature, and to the significant or sacred”^9^ Spirituality has been found to play vital roles in health through disease detection and treatment compliance, beliefs that conflict with medical care, medical decision making and spiritual struggles that create stress and impair health outcome.^10^ Specifically, it has been noted as an essential factor influencing the health-related quality of life of chronically ill patients due to its integration into positive thoughts and health behaviours, as well as healthy coping styles which alleviate the perceived overwhelming persistent situation.^11,12^

Spirituality has been specifically associated with good blood pressure control in hypertensive patients.^13^ The importance of addressing spirituality in hypertension like in other chronic diseases is indicated in several studies.^8,12,14^ Spirituality has been shown to be important to hypertensive patients, and some patients have indicated that they use their spirituality for coping and disease management.^15^ In a study conducted by Kretchy et al.,^8^ findings revealed that participants with a high level of spirituality and religious participation had significantly lower diastolic and systolic blood pressure than their counterparts with a low level of spirituality (77.8 vs. 84.7 mmHg and 137.2 vs. 149.5 mmHg, respectively) after adjusting for demographic, sociocultural, and psychological variables.^16^ Studies have also revealed that people who engaged in spiritual activities had lower levels of cortisol (a biological marker of stress), lipid profiles, glucose control and immune function.^16-18^

The relationship between spirituality and hypertension has been studied by several researchers in Africa. Some of them revealed a positive correlation between spirituality, medication adherence and blood pressure control while others did not.^8^ However; there is a paucity of studies on the relationship between spirituality and hypertension in Nigeria.

This study is designed to determine the role of spirituality in the control of blood pressure among adult hypertensive patients. It also seeks to answer the following research questions:

1. What is the level of spirituality among adult hypertensive patients?
2. What is the level of blood pressure control among the respondents?
3. What is the relationship between the level of spirituality and the socio-demographic characteristics of respondents?
4. What is the relationship between the level of spirituality and blood pressure control?

## MATERIALS AND METHOD

### STUDY AREA/STUDY DESIGN/POPULATION

The hospital-based descriptive cross-sectional study was conducted among adult hypertensive patients aged 18 years and above attending the Family Medicine Clinic of Federal Teaching Hospital, Ido-Ekiti (FETHI) in rural Southwest Nigeria. The study was conducted over sixteen (16) weeks from April to July 2017 during which a total of 1,986 hypertensive patients were encountered.

### SAMPLE SIZE DETERMINATION

The sample size was calculated using the formula for estimating proportions in a cross-sectional study: n = z^2^ pq / d^2^ (For population >10,000)^19^ and nf = n/1+ (n/N) (For population <10,000).^19^ Where:

n = the minimum sample size when the population is more than 10,000.

nf = the minimum sample size when the population is less than 10,000.

N = the estimate of study population size in a given year, i.e. adult hypertensive patients who attended the Family Medicine Clinic of Federal Teaching Hospital in 2015. (N= 4,800).

z_α_ = the level of confidence (The standard normal deviation set at 1.96 which corresponds to the 95% confidence level obtained from the standard statistical table of normal distribution.

p = Estimated prevalence of medication adherence among hypertensive patients in a given population, (using 25.6%, which was the prevalence of medication adherence among hypertensive patients at a tertiary hospital, in South-Western Nigeria).^20^

q = 1 – p.

d = Degree of accuracy desired usually set at 0.05

The minimum sample size was 276 respondents. However, to allow for non-responses during recruitment, a non-response value of 10% (28) was added to the minimum sample size (nf). This gave a minimum sample size of 304 respondents. Hence, 304 respondents were used for the study. A sampling frame was built, by sorting out the case files of hypertensive patients attending the clinic for the day.

The selected hypertensive patients attending the clinic for the day were selected from the general patient pool, and directed to another convenient room within the family medicine complex. The potential benefits of the study were explained to them and verbal consents were obtained. The consenting patients were subjected to the inclusion and exclusion criteria. The number of consenting hypertensive patients who met the inclusion and exclusion criteria daily were between 19 and 21 out of an average of 25 participants and a total of 1,628 hypertensive patients out of the 1,986 encountered patients both consented and met the inclusion and exclusion criteria during the 16 weeks of the study.

Participants who met the inclusion and exclusion criteria on each clinic day were assigned number from 1 up to the last number for the day.

The sampling interval (K) was calculated using the formula:

K = N/n^19^

(Where: N = total number of patients, n = calculated minimum sample size and K = sampling interval).

Hence K = 1628/304 = 5.36 which is approximately 5.

Systematic random sampling technique was then used to select the participants on each clinic day; hence, every 6^th^ consenting hypertensive patient who met the inclusion criteria was enrolled for this research. Therefore, an average of four (4) hypertensive patients was recruited per day.

To ensure that a patient was not selected twice, a register for all patients that participated in the study was opened, using their hospital numbers and a colour tag on their file jackets.

### INCLUSION CRITERIA

1. All consenting adult hypertensive patients aged 18 years and above.
2. Patients who have been diagnosed and on treatments for hypertension for at least 3 months in the clinic and had recorded a minimum of three (3) consecutive clinic visits in the last six months before recruitment to ensure familiarity with diagnosis and prescribed treatment modalities.

### EXCLUSION CRITERIA

1. Patients with co-morbidity (Chronic Kidney Disease, Diabetes)
2. Critically ill patient as at the time of the study (Hypertensive emergency, Congestive cardiac failure, Cerebrovascular accident).
3. Pregnant hypertensive patients because they belong entirely to another class of hypertension.

### DATA COLLECTION AND INSTRUMENTS

Socio-demographic variables were obtained using a semi-structured interviewer-administered questionnaire Clinical parameters (including BP) of respondents were measured, recorded and graded by the researcher.

Assessment of patients’ level of Spirituality was done using the Spiritual perspective scale (SPS). It is a 10-item tool designed to measure the extent to which individuals hold certain spiritual beliefs and engage in spiritually related behaviours.^16,148^ It was scored using the mean.^8,21^ The mean spirituality score for this study was 40.9±15.2. The scoring and determination of the level of spirituality are found below (section 3.9.3.3.1).^21^ Scores within 10-35 was indicated a low level of spirituality and those ranging between 36-60 indicated a high level of spirituality. Using Cronbach’s alpha, reliability has consistently been rated above 0.90 and average inter-item correlations are 0.878.^8,21,22^ The SPS has been used successfully in prior African studies.^8,22^

### Scoring and Determination of the Level of Spirituality^23^

Ten items are measuring the beliefs and behaviours of respondents on a 6 point Likert scale. The minimum and maximum obtainable scores were 10 and 60 respectively.

Range = minimum possible score – maximum possible score

Number of expected outcome = Respondents’ level of spirituality dichotomised into high or low .i.e. 2 divisions

For this study, respondents’ level of spirituality is dichotomised into high or low. This gives a range of 60 – 10 = 50

Interval = <u>Range</u> (50)

The number of expected outcomes (2). Interval is 25

Therefore

10 to 35 (minimum score + interval) is a low level of spirituality 36 to 60 is a high level of spirituality

Assessment of Blood Pressure was measured with an Accoson® brand mercury sphygmomanometer. For this study, a cut-off level of both < 140mmHg and < 90mmHg for SBP and DBP respectively were considered Controlled BP. Uncontrolled BP was considered when the SBP and DBP were ≥ 140mmHg and ≥ 90mmHg respectively using the recommendation of eight Report of the Joint National Committee on Prevention, Detection, Evaluation, and Treatment of High Blood Pressure (JNC-VIII) criteria. ^24^

### DATA ANALYSIS

All data collected were analyzed using the Statistical Package for Social Sciences (SPSS) for Windows software version 20 (SPSS Inc., Chicago, IL, USA).^25^ The data were presented in tabular form, graphs and charts as appropriate. Frequency tables and charts were generated for relevant variables. Means, standard deviations, and percentages were determined as appropriate. The means, median and standard deviation were calculated for continuous variables while categorical variables were analyzed using proportions. The Pearson’s chi-square test was used to assess the bivariate association between spirituality with the respondents’ socio-demographic characteristics and blood pressure control and the Fisher’s exact test was used in analyzing variables with cells less than 5 counts. Binary logistic regression analysis was used to assess the association between the independent variable (spirituality) and dependent variable (blood pressure control) while adjusting for the socio-demographic characteristics. A P-value of equal or less than 0.05 was taken to be statistically significant.

### ETHICAL CLEARANCE AND CONSENT

Ethical approval was obtained from the Ethics and Research Committee of Federal Teaching Hospital, Ido-Ekiti (Protocol Number: ERC/2016/02/25/08A). Informed verbal consent was obtained from the willing participants. To maintain confidentiality, no names appeared on the questionnaires, but only numbers were used as identifiers. The reporting of this study conforms to the strengthening the Reporting of observational studies in Epidemiology (STROBE) statement^26^. All patients’ details have been de-identified.

## RESULTS

### 4.1 Characteristics of Respondents

A total of 304 respondents participated in the study. The respondent’s ages ranged between 18 and 82 years, their mean age was 61.1±11.1 years and the majority (57.9%) were older adults (65-82 years). The majority of respondents were female (53.6%) and were married (76.0%), Christians (78.6%) and rural dwellers (64.5%) from the Yoruba tribe (74.7%). Most respondents were traders (37.5%) who had a primary level of education (31.9%) and earned incomes below the poverty line (57.7%). The details of the respondent’s socio-demographic characteristics were as shown in Table 1 below.

**Table 1:** Socio-demographic Characteristics of Respondents.

| **Variable** | **Frequency**<br>**(n = 304)** | **Percentage**<br>**(%)** |
| --- | --- | --- |
| **Age (years)** | ***61.1 ± 11.1*** |  |
| Young Adult (18-35) | 6 | 2.0 |
| Middle-Aged Adult (36-64) | 122 | 40.1 |
| Older Adult (65-82) | 176 | 57.9 |
| **Sex** |  |  |
| Male | 141 | 46.4 |
| Female | 163 | 53.6 |
| **Marital Status** |  |  |
| Single | 10 | 3.3 |
| Married | 231 | 76.0 |
| Divorced/Separated | 43 | 14.1 |
| Widowed | 20 | 6.6 |
| **Religion** |  |  |
| Christianity | 239 | 78.6 |
| Islam | 59 | 19.4 |
| Traditionalist | 6 | 2.0 |
| **Domicile** |  |  |
| Urban | 108 | 35.5 |
| Rural | 196 | 64.5 |
| **Ethnicity** |  |  |
| Yoruba | 227 | 74.7 |
| Igbo | 46 | 15.1 |
| Hausa | 26 | 8.6 |
| Others | 5 | 1.6 |
| **Educational Level** |  |  |
| None | 38 | 12.5 |
| Primary | 97 | 31.9 |
| Secondary | 63 | 20.7 |
| Tertiary | 88 | 29.0 |
| Post Graduate | 18 | 5.9 |
| **Occupation** |  |  |
| Professionals | 61 | 20.1 |
| Trading | 114 | 37.5 |
| Farming | 67 | 22.0 |
| Retiree | 32 | 10.5 |
| Self – Employed | 30 | 9.9 |
| **Income\*** |  |  |
| < Poverty Line (<₦20,520) | 175 | 57.7 |
| ≥Poverty Line (≥₦20,520) | 129 | 42.3 |

*₦20,520 is the expected minimum income per month equivalent of \$1.90 per day at \$1 equals to ₦360 (as of July 5, 2017)<sup>27</sup>*

Table 2 showed the Spiritual perspective scale based on the spiritual behaviours and beliefs of respondents. Spiritual behaviours that contributed significantly to the high level of spirituality in this study were: private prayers/meditations (χ^2^ = 3.42, SD = 0.97) and spiritual discussions with family and friends (χ^2^ = 3.19, SD = 0.92), while Spiritual beliefs that contributed significantly to the high level of spirituality in this study were: forgiveness (χ^2^ = 3.27, SD = 0.74), seeking spiritual guidance before making decisions (χ^2^ = 3.26, SD = 0.87) and regarding spirituality as a means of finding live meaning and solving life’s puzzle (χ^2^ = 3.26, SD = 0.64).

**Table 2a:** Spiritual Perspective Scale.

| S/N | SPS (Spiritual Behaviours) | Not at all | Less than once a year | About once a year | About once a month | About once a week | About once a day | $\Sigma F$ | Mean | Standard Deviation |
| --- | --- | --- | --- | --- | --- | --- | --- | --- | --- | --- |
| 1 | In talking with family and friends, how often do you mention spirituality? | 4 | 10 | 16 | 35 | 102 | 137 | 304 | 3.19 | 0.92 |
| 2 | How often do you share with others the problems and joys of living according to your spiritual beliefs? | 10 | 11 | 10 | 54 | 102 | 117 | 304 | 3.08 | 1.10 |
| 3 | How often do you read spiritually-related material? | 14 | 15 | 20 | 50 | 92 | 113 | 304 | 2.95 | 1.11 |
| 4 | How often do you engage in private prayer and meditation? | 9 | 10 | 6 | 36 | 48 | 195 | 304 | 3.42 | 0.97 |

**Table 2b:**
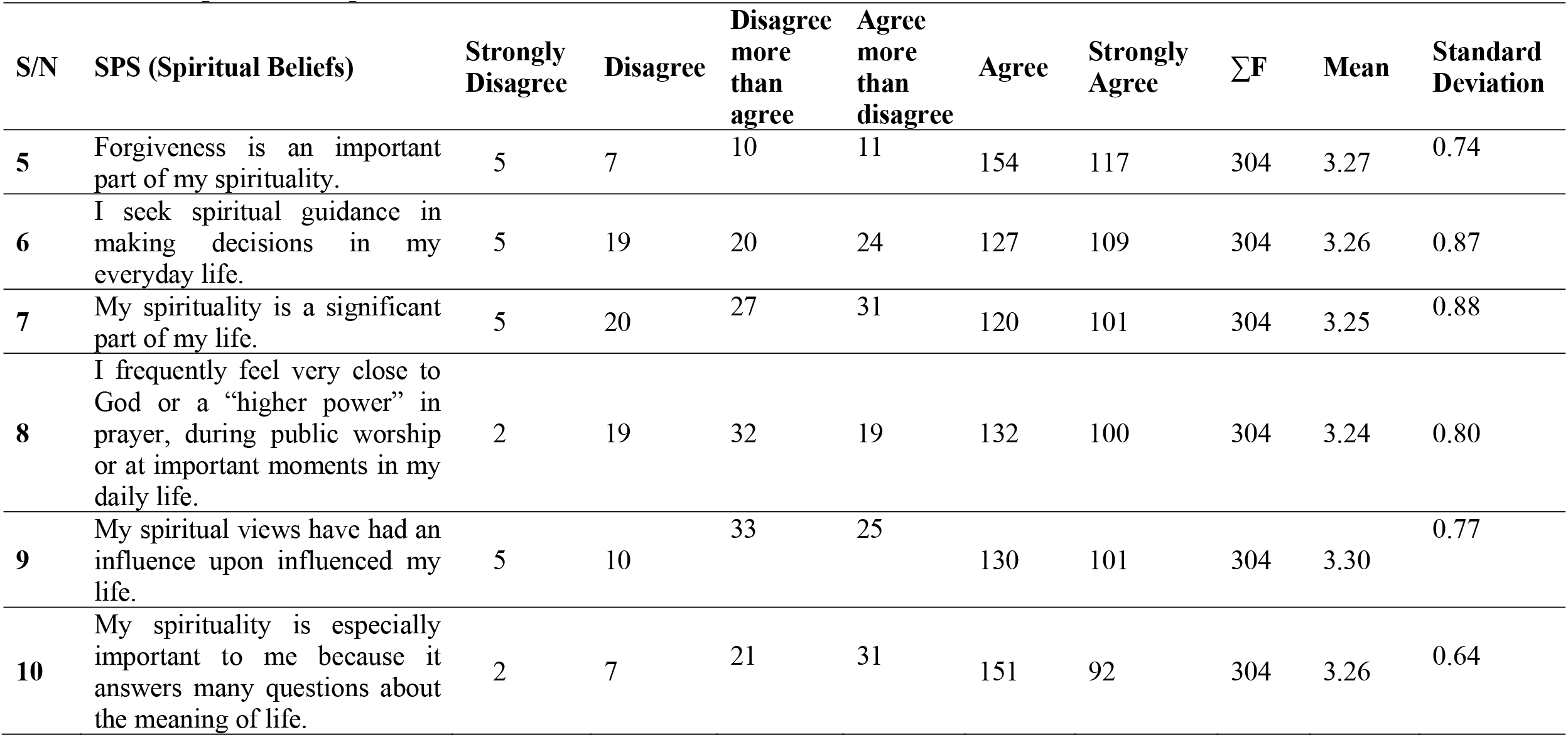
Spiritual Perspective Scale.

| S/N | SPS (Spiritual Beliefs) | Strongly Disagree | Disagree | Disagree more than agree | Agree more than disagree | Agree | Strongly Agree | $\Sigma F$ | Mean | Standard Deviation |
| --- | --- | --- | --- | --- | --- | --- | --- | --- | --- | --- |
| 5 | Forgiveness is an important part of my spirituality. | 5 | 7 | 10 | 11 | 154 | 117 | 304 | 3.27 | 0.74 |
| 6 | I seek spiritual guidance in making decisions in my everyday life. | 5 | 19 | 20 | 24 | 127 | 109 | 304 | 3.26 | 0.87 |
| 7 | My spirituality is a significant part of my life. | 5 | 20 | 27 | 31 | 120 | 101 | 304 | 3.25 | 0.88 |
| 8 | I frequently feel very close to God or a “higher power” in prayer, during public worship or at important moments in my daily life. | 2 | 19 | 32 | 19 | 132 | 100 | 304 | 3.24 | 0.80 |
| 9 | My spiritual views have had an influence upon influenced my life. | 5 | 10 | 33 | 25 | 130 | 101 | 304 | 3.30 | 0.77 |
| 10 | My spirituality is especially important to me because it answers many questions about the meaning of life. | 2 | 7 | 21 | 31 | 151 | 92 | 304 | 3.26 | 0.64 |

Table 3 showed the level of spirituality among the respondents. More than half (52.6%) of the respondents had a high level of spirituality while 47.4% had a low level of spirituality. The mean spirituality score obtained was 40.9±15.2.

**Table 3:** Assessment of the Level of Spirituality among the Respondents.

| Variable | Frequency<br>n= 304 | Percentage<br>(%) |
| --- | --- | --- |
| **Spirituality Perspective Scale** |  |  |
| Low (10-35) | 144 | 47.4 |
| High (36-60) | 160 | 52.6 |
| **Total** | **304** | **100.0** |
| *Mean Spirituality Score ± SD* | *40.9 ± 15.2* |  |

Figure 2. : A bar chart showing the level of blood pressure control among the respondents. More than two-thirds (67.1%) of the respondents had controlled blood pressure while 32.9% had uncontrolled blood pressure

**Figure 1:**
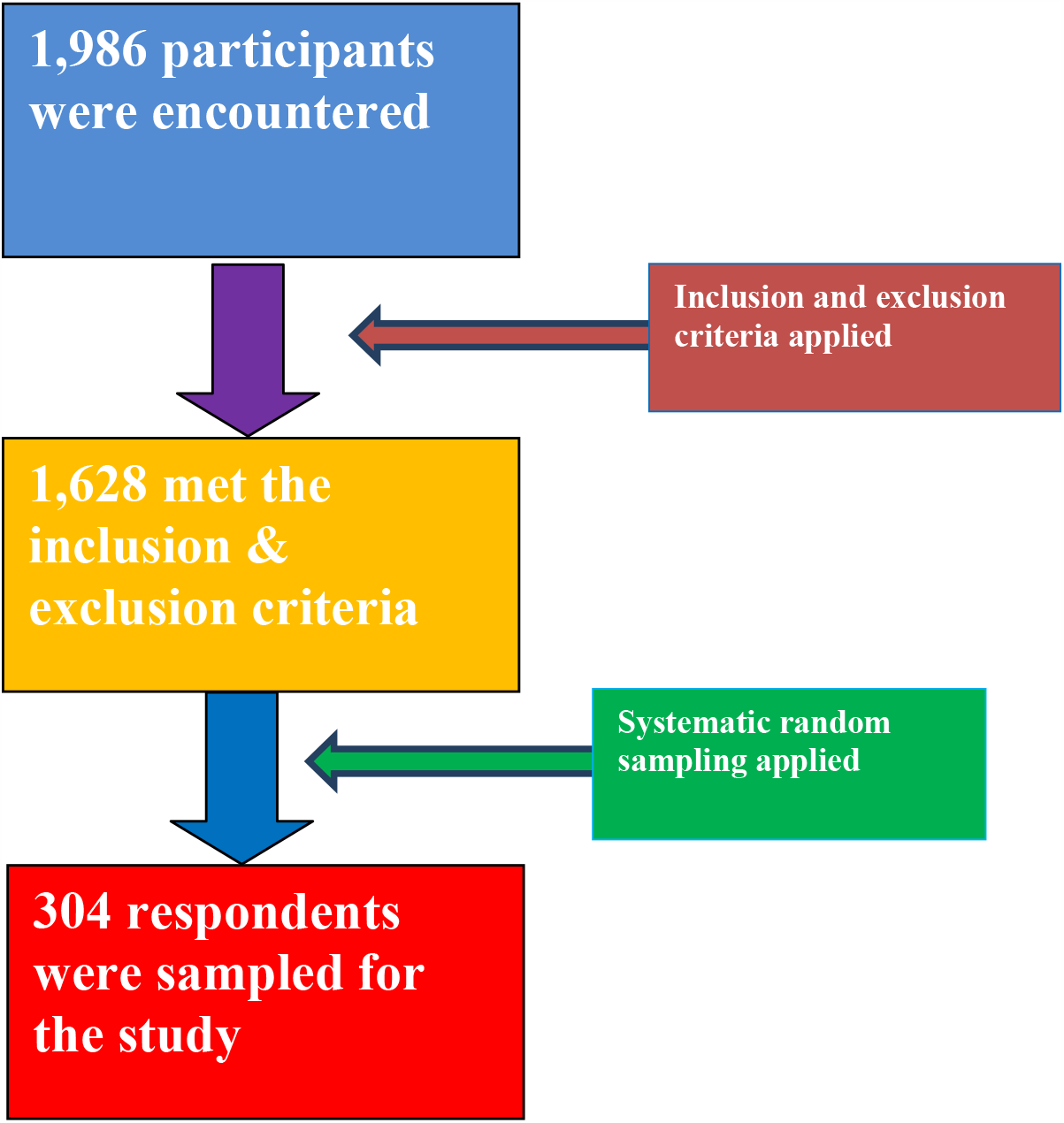
Study flowchart.

**Figure 2:**
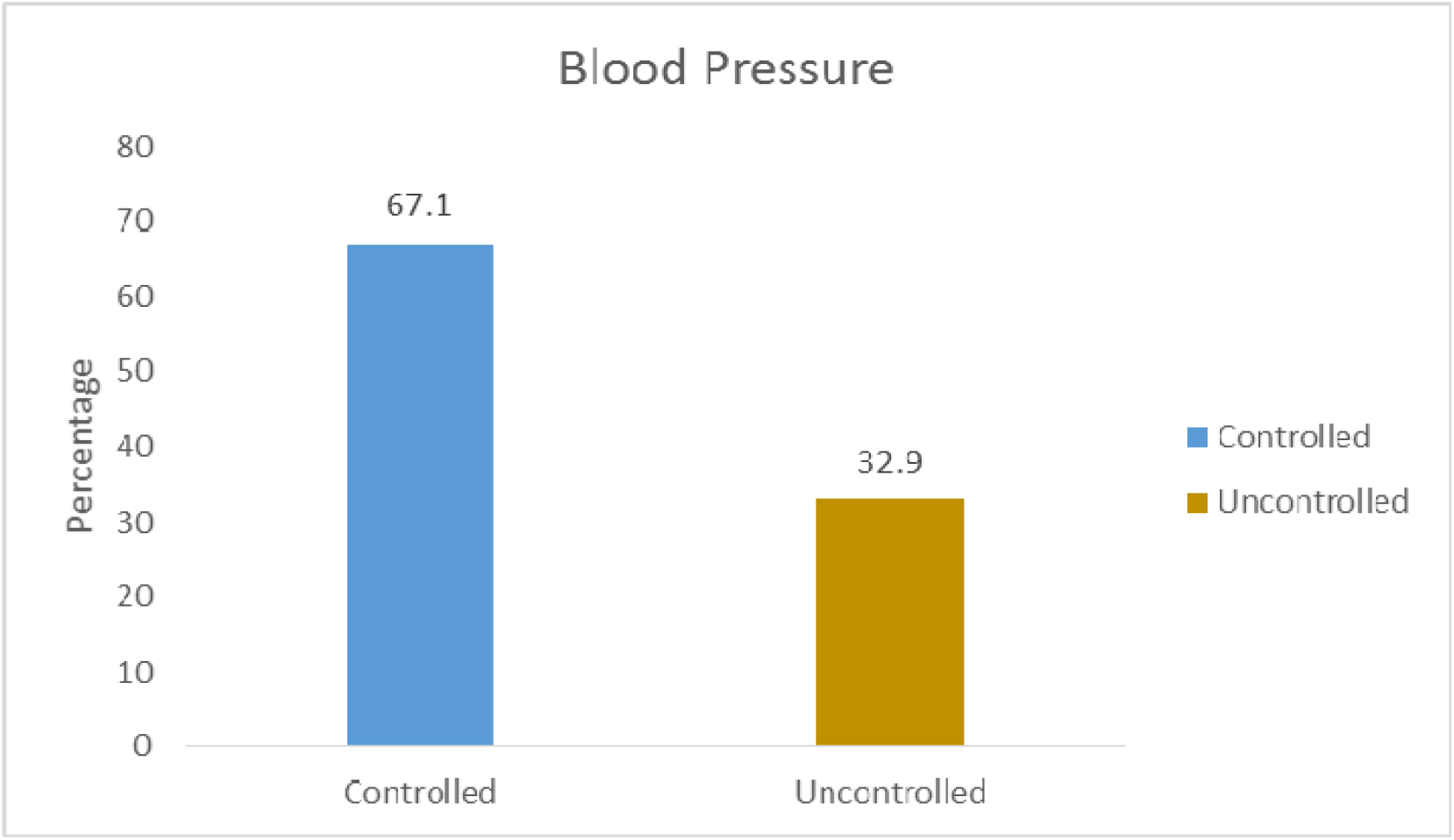
A graphical representation of the level of Blood Pressure Control among Respondents.

Table 4 showed the relationship between spirituality and blood pressure among the respondents. It showed that Blood pressure control improves with increasing spirituality. More than two-thirds (70.1%) of those with a high level of spirituality also had good blood pressure control. The relationship was statistically significant as p<0.05.

**Table 4:**
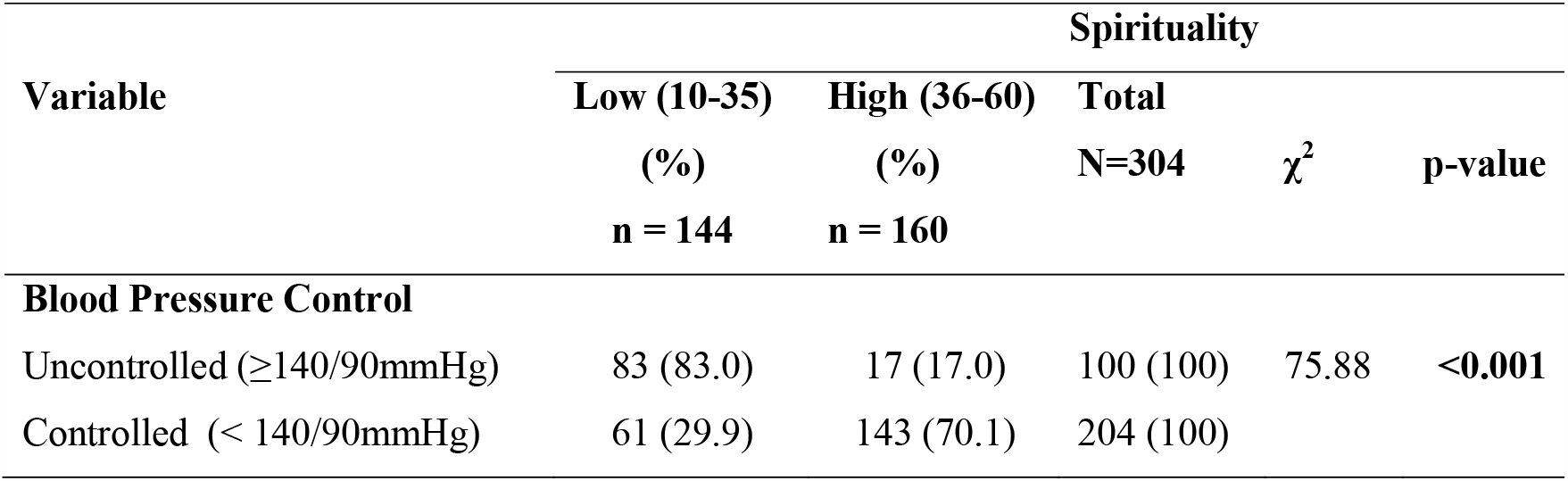
Relationships between Blood Pressure Control and Spirituality among the Respondents.

Table 5 showed the relationships between the socio-demographic characteristics of respondents and their level of spirituality. Older adult (64.4%), female (67.5%), married (58.4%), Christian (56.5%), Yoruba (63.0%) and urban dwelling (55.1%) respondents had high level of spirituality. Also, respondents that had a tertiary level of education (70.4%) who are retired (66.7%) and earned below the poverty line (75.4%) had a high level of spirituality. All variables were statistically significant with p<0.05 except the area of domicile.

**Table 5:** Relationships between the Level of Spirituality and Sociodemographic Characteristics of Respondents.

| Variable | Spirituality | | | $\chi^2$ | p-value |
| --- | --- | --- | --- | --- | --- |
|  | Low (%)<br>n = 144 | High (%)<br>n = 160 | Total<br>N=304 |  |  |
| **Age (years)** |  |  |  |  |  |
| Young Adult (18-35) | 4 (66.7) | 2 (33.3) | 6 (100) |  | **0.004<sup>‡</sup>** |
| Middle-Aged Adult (36-64) | 77 (63.6) | 44 (36.4) | 121 (100) |  |  |
| Older Adult (65-82) | 63 (35.6) | 114 (64.4) | 177 (100) |  |  |
| **Sex** |  |  |  |  |  |
| Male | 91 (64.5) | 50 (35.5) | 141 (100) | 31.09 | **<0.001** |
| Female | 53 (32.5) | 110 (67.5) | 163 (100) |  |  |
| **Marital Status** |  |  |  |  |  |
| Single | 10 (100.0) | 0 (0.0) | 10 (100) |  | **0.001<sup>‡</sup>** |
| Married | 96 (41.6) | 135 (58.4) | 231 (100) |  |  |
| Divorced | 15 (75.0) | 5 (25.0) | 20 (100) |  |  |
| Separated | 14 (60.9) | 9 (39.1) | 23 (100) |  |  |
| Widowed | 9 (45.0) | 11 (55.0) | 20 (100) |  |  |
| **Religion** |  |  |  |  |  |
| Christianity | 104 (43.5) | 135 (56.5) | 239 (100) |  | **0.018<sup>‡</sup>** |
| Islam | 37 (62.7) | 22 (37.3) | 59 (100) |  |  |
| Traditionalist | 4 (66.7) | 2 (33.3) | 6 (100) |  |  |
| **Domicile** |  |  |  |  |  |
| Urban | 88 (44.9) | 108 (55.1) | 196 (100) | 1.35 | 0.245 |
| Rural | 56 (51.9) | 52 (48.1) | 108 (100) |  |  |
| **Ethnicity** |  |  |  |  |  |
| Yoruba | 84 (37.0) | 143 (63.0) | 227 (100) |  | **<0.001<sup>‡</sup>** |
| Igbo | 39 (84.8) | 7 (15.2) | 46 (100) |  |  |
| Hausa | 18 (69.2) | 8 (30.8) | 26 (100) |  |  |
| Others | 3 (60.0) | 2 (40.0) | 5 (100) |  |  |
| **Educational Status** |  |  |  |  |  |
| None | 24 (63.2) | 14 (36.8) | 38 (100) | 21.57 | **<0.001** |
| Primary | 57 (58.8) | 40 (41.2) | 97 (100) |  |  |
| Secondary | 31 (49.2) | 32 (50.8) | 63 (100) |  |  |
| Tertiary | 26 (29.6) | 62 (70.4) | 88 (100) |  |  |
| Post Graduate | 6 (33.3) | 12 (66.7) | 18 (100) |  |  |
| **Occupation** |  |  |  |  |  |
| Professionals | 24 (39.3) | 37 (60.7) | 61 (100) | 13.43 | **0.009** |
| Trading | 49 (43.0) | 65 (57.0) | 114 (100) |  |  |
| Farming | 43 (64.2) | 24 (35.8) | 67 (100) |  |  |
| Self Employed | 18 (56.3) | 14 (43.7) | 32 (100) |  |  |
| Retired | 10 (33.3) | 20 (66.7) | 30 (100) |  |  |
| **Income** |  |  |  |  |  |
| <Poverty Line | 43 (24.6) | 132 (75.4) | 175 (100) | 85.96 | **<0.001** |
| ≥Poverty Line | 101 (78.3) | 28 (21.7) | 129 (100) |  |  |

<sup>‡</sup> = Fisher's Exact Test

Table 6 explains how some socio-demographic characteristics predicted the level of spirituality. Respondents who were middle-aged adults (1.54 times), Married (7.54 times), Christians (3.38 times), Yoruba (2.23 times), Postgraduate (2.70 times), Professionals (3.54), Less than 5 years on antihypertensive drug (2.72), Controlled blood pressure (4.76 times) and were on 2 drugs or less (2.80 times) were more likely to have a high level of spirituality. However, respondents who were male gender (0.71 times), farmers (0.78) and earned income below the poverty line (0.74 times) were less likely to have high medication adherence.

**Table 6:** Logistics Regression for predictors of Spirituality among the Respondents.

| Variable |  | OR | 95% CI |  | p-value |
| --- | --- | --- | --- | --- | --- |
|  |  |  | Lower | Upper |  |
| **Age group** | Young Adult (18-35) | 0.54 | 0.53 | 2.68 | 0.051 |
|  | Middle-Aged Adult (36-65) | 1.54 | 1.05 | 8.42 | **0.003** |
|  | Older Adult (>65) | 1.0 (RC) |  |  |  |
| **Sex** | Male | 0.71 | 0.04 | 0.90 | **0.021** |
|  | Female | 1.0 (RC) |  |  |  |
| **Religion** | Christianity | 3.38 | 1.22 | 10.81 | **0.004** |
|  | Islam | 1.11 | 1.08 | 3.29 | **0.031** |
|  | Traditionalist | 1.0 (RC) |  |  |  |
| **Ethnicity** | Yoruba | 2.23 | 1.19 | 9.41 | **0.011** |
|  | Igbo | 1.53 | 1.04 | 2.92 | **0.016** |
|  | Hausa | 1.10 | 0.03 | 3.05 | 0.062 |
|  | Others | 1.0 (RC) |  |  |  |
| **Education** | None | 1.0 (RC) |  |  |  |
|  | Primary | 1.55 | 0.13 | 6.93 | 0.051 |
|  | Secondary | 1.11 | 1.04 | 7.42 | **0.012** |
|  | Tertiary | 2.13 | 1.67 | 10.54 | **0.037** |
|  | Postgraduate | 2.70 | 1.08 | 9.93 | **0.030** |
| **Occupation** | Professionals | 3.54 | 1.04 | 9.47 | **0.022** |
|  | Trading | 1.21 | 1.02 | 6.21 | **0.004** |
|  | Farming | 0.78 | 0.09 | 0.99 | **0.016** |
|  | Self – Employed | 3.02 | 1.98 | 9.83 | **0.039** |
|  | Retired | 1.0 (RC) |  |  |  |
| **Income** | < Poverty Line | 0.74 | 0.10 | 0.97 | **0.006** |
|  | >Poverty Line | 1.0 (RC) |  |  |  |

## DISCUSSION

### SOCIO-DEMOGRAPHIC CHARACTERISTICS OF RESPONDENTS

In Table 1, the socio-demographic profile of the respondents showed that the mean age of patients with hypertension in this study was 61.1±11.1 years with the majority of the respondents being above 65 years. This was higher than 50.9±9.1 years reported in a similar hospital-based study in Port-Harcourt by Akpa *et al*^28^ and 50.6±1.5 years reported in a study by Azinge *et al* in Lagos^29^. It was however lower than the mean age of 63.1±5.7 years found in an international survey.^30^ The reason for the higher preponderance of hypertension in the older age group found in this study compared to other studies might be because this study was done in a rural population as this usually consists of older age group due to rural to urban migration of the young age groups. However, Ido-Ekiti town is a non-industrialised semi-urban setting surrounded by rural settlements.

The majority of the respondents {163 (53.6%)} in this study were females. This aligned with the findings of Iloh et al. (65.2%)^31^ who also reported a higher prevalence of hypertension in females. It is worthy of note that women are protected from most cardiovascular events compared to men, until after menopause, during which women are at increased risk of cardiovascular complications compared to premenopausal women.^32,33^ Gender differences in blood pressure is inconsistent in many population studies, while some studies quoted a higher prevalence of hypertension among males,^34,35^ others reported no significant gender difference.^28,36^ These inconsistencies could be as a result of different age populations or the methodology used in these studies.

In this study about two-thirds {231 (76%)} of all the respondents were married. This was similar to findings by Marfo et al.^37^ who reported 74% married hypertensive patients.^37^ However, findings in this study was higher than the 66.4% reported in Ibadan by Ekore *et al*.^30^ This difference may be due to the variation in marriage culture in different states and regions among the study populations in Nigeria. The percentage of respondents who did not have any formal education was 12.5%. It could be inferred that 87.5% had formal education in this study. This might be due to the study area which is located in the South-Western part of the country where there is a high literacy level which might influence patients’ health-seeking behaviour in terms of them presenting in the hospital for the management of their disease. This study confirms the rating of Ekiti State as an educationally advantaged state. The reported level of literacy was much higher in a hospital-based study done among young adults in Ibadan, South-western Nigeria where about 79.1% of respondents had a minimum of secondary school education^36^ and 40.0% reported in Lagos among market women.^,38^ These differences may be due to the different study populations among whom the studies were conducted.

### LEVEL OF SPIRITUALITY AMONG RESPONDENTS

In Table 3, this study showed that the majority of the respondents (52.6%) had a high level of spirituality. The finding of the majority of the respondents being highly spiritual in this study may not be unconnected with the cultural practices which were inherent in Nigeria. This was similar to a study by Ibraheem et al^39^ in the relationship between self-reported health status and spirituality among adult patients in Ibadan, Nigeria who found a 51% level of spirituality.^39^ In contrast, Jeri and Lynda reported 100% spirituality level among Nigerians living in America.^40^ This is higher than that obtained in the present study, and the difference may be due to the different study populations among whom the studies were conducted and the method used in the study.

In Table 2. Spiritual behaviours that contributed significantly to the high level of spirituality in this study according to Table 3 were: private prayers/meditations (χ^2^ = 3.42, SD = 0.97) and spiritual discussions with family and friends (χ^2^ = 3.19, SD = 0.92). Also, Spiritual beliefs that contributed significantly to the high level of spirituality in this study were: forgiveness (χ^2^ = 3.27, SD = 0.74), seeking spiritual guidance before making decisions (χ^2^ = 3.26, SD = 0.87) and regarding spirituality as a means of finding meaning to live and solving life’s puzzle (χ^2^ = 3.26, SD = 0.64).

### 5.8. RELATIONSHIP BETWEEN SPIRITUALITY AND BLOOD PRESSURE CONTROL

In Table 4, this study has shown a high spirituality level of 52.6% and a blood pressure control level of 67.1%. The Chi-square test showed a significant relationship between them with p<0.001. This is similar to the finding in a study done on spirituality and blood pressure control by Kretchy et al.,^8^ in Umuahia, Nigeria, which showed participants with a high level of spirituality and religious participation had significantly lower diastolic and systolic blood pressure than their counterparts with a low level of spirituality (77.8 vs. 84.7 mmHg and 137.2 vs. 149.5 mmHg, respectively) after adjusting for demographic, sociocultural, and psychological variables.^8^

However, the findings in this study are higher than what was observed by Quingtao et al.^41^ in a study conducted in china who found that 53.4% of those who had a high level of spirituality also had good blood pressure control.

Also, logistic regression done in this study according to Table 6, showed that for a single unit increase in the level of spirituality, hypertensive patients were 4.76 times more likely to achieve good blood pressure control (OR= 4.76, 95%CI = 1.05-14.99, p<0.001). Hence, a high level of spirituality is a significant predictor of good blood pressure control among hypertensive patients.

#### RELATIONSHIP BETWEEN SPIRITUALITY AND SOCIO-DEMOGRAPHIC CHARACTERISTICS OF RESPONDENTS

In Table 5, this study found that spirituality levels increased with age and specifically, more than half (64.4%) of the older aged group has a higher level of spirituality. This could be as a result of the fact that, as people get older they tend to find solace and meaning to life in the transcendent and hence tend to have more inclination towards spirituality.^42^ The above finding is in agreement with hospital-based research conducted by Ibraheem et al (83.9%)^39^ who also reported an increased level of spirituality with age.

This study established the fact that women have higher spirituality than men (67.5%). This is possibly due to the vulnerability of women in this part of the world and women tend to find solace in their spiritual and religious practices. This is also similar to what Jeri et al. (63%)^40^ found in his study.

In this study, a higher spirituality score was found among the respondents who were married (58.4%) when compared with the respondents who were single or divorced and this difference was statistically significant (p=0.001). This might be due to the possible encouragement by the spouse towards spirituality, which is not found among the single and the divorced. This trend was in agreement with the finding of Ibraheem et al.^39^ in his study on the relationship between self-reported health status and spirituality among adult patients attending the General outpatient clinic of a tertiary hospital in Ibadan where he reported a higher spirituality level among married people (68%) compared with those who were not married.^39^

It was observed in this study that spirituality increased with an increase in the level of literacy and this was statistically significant (p<0.001). This might be due to the high literacy level found in a large proportion of the respondents in this study. This is different from the previous finding of Silva et al.^43^ who reported that people of low socio-economic class tend to be more spiritual and that the prevalence of spiritual commitment tends to drop off among higher-income categories.^43^

### 5.10. CONCLUSION

This study has shown the relationship between spirituality and blood pressure control among adult hypertensive patients in rural Southwestern, Nigeria.

It had shown 52.6% high spirituality level and 67.1% good blood pressure control.

The study found that more than two-thirds of respondents with a high level of spirituality had good blood pressure control, while more than three-quarters of respondents with a low level of spirituality had poor blood pressure control. These relationships were statistically significant.

These associations provide direction for future studies that will aid in understanding how health professionals can use this information to provide culturally sensitive and patient-centred care that will improve blood pressure control among patients with hypertension. Several studies emphasized the importance of spirituality in overall health and some behaviour of chronically ill persons; however, spirituality has been largely overlooked in the study of blood pressure control. This study addresses this gap in the literature regarding the association between spirituality and blood pressure control in adult hypertensive patients.

### 5.11. RECOMMENDATIONS

The study has highlighted the significance of the relationship between spirituality and blood pressure control among patients with hypertension; hence, Physicians should take appropriate spiritual history while managing patients with hypertension to identify how spirituality can positively or negatively affect blood pressure management and control.

Also, the Physicians should encourage the health facilities to have chaplains and Imams that patients can be referred to for spiritual assistance when needed.

It is also important that the Physicians understand their limits in the process of integrating spirituality into their patient’s care.

### 5.12 LIMITATIONS OF THE STUDY

1. The study is a cross-sectional study done in a tertiary health institution, the findings may not apply to the general population.
2. Subjects’ ambulatory or home blood pressure readings in out-of-clinic settings were not monitored in this study, and thus the determination of optimal blood pressure control was based on clinic-based measurement alone instead of an average over a period of time.

## Data Availability

All data are available in the manuscript

## Acknowledgements

The authors would like to appreciate nurses and resident doctors of family medicine department and the management of FETHI where the study was conducted.

## Availability of data and materials

The datasets for this study would be made available from the correspondence author on a reasonable request.

## Authors’ contribution

1. AO - Conceptualization of the study, designed the study protocol, data acquisition and analysis and drafted the initial manuscript
2. PO - Literature review, data analysis and review of manuscript for intellectual content.
3. OMS - Critically revised the protocol for methodological and intellectual content.
4. AOI. Literature review, review of manuscript for intellectual content.

All authors have read and approved the final version of the manuscript prior to submission.

